# The impact of temperature on the transmission potential and virulence of COVID-19 in Tokyo, Japan

**DOI:** 10.1101/2021.06.15.21258529

**Authors:** Lisa Yamasaki, Hiroaki Murayama, Masahiro Hashizume

## Abstract

**Background:** Assessing the impact of temperature on COVID-19 epidemiology is critical for implementing non-pharmaceutical interventions. However, few studies have accounted for the nature of contagious diseases, i.e., their dependent happenings.

**Aim:** We aimed to quantify the impact of temperature on the transmissibility and virulence of COVID-19 in Tokyo, Japan. We employed two epidemiological measurements of transmissibility and severity: the effective reproduction number (*R*_*t*_) and case fatality risk (CFR).

**Methods:** We used empirical surveillance data and meteorological data in Tokyo to estimate the *R*_*t*_ and time-delay adjusted CFR and to subsequently assess the nonlinear and delay effect of temperature on *R*_*t*_ and time-delay adjusted CFR.

**Results:** For *R*_*t*_ at low temperatures, the cumulative relative risk (RR) at first temperature percentile (3.3°C) was 1.3 (95% confidence interval (CI): 1.1-1.7). As for the virulence to humans, moderate cold temperatures were associated with higher CFR, and CFR also increased as the temperature rose. The cumulative RR at the 10^th^ and 99^th^ percentiles of temperature (5.8°C and 30.8°C) for CFR were 3.5 (95%CI: 1.3-10) and 6.4 (95%CI: 4.1-10.1).

**Conclusions:** This study provided information on the effects of temperature on the COVID-19 epidemiology using *R*_*t*_ and time-delay adjusted CFR. Our results suggest the importance to take precautions to avoid infection in both cold and warm seasons to avoid severe cases of COVID-19. The results and proposed framework will also help in assessing possible seasonal course of COVID-19 in the future.

## Introduction

The COVID-19 pandemic has imposed significant health and economic burdens all over the world. A better understanding of the factors affecting the COVID-19 epidemic is critical to the design of tailored public health and social measures (PHSMs), e.g., travel restrictions, school closures, cancellation of public events and gatherings, etc. and much attention has been given to the impact of meteorological factors on the COVID-19 transmissibility and severity.

Over the last couple of decades, important factors related to the transmission of viral respiratory diseases have been investigated such as the highly predictable seasonal pattern of influenza epidemics [1]. These epidemiological studies are supported by laboratory evidence that low temperature and/or humidity improve the stability of influenza virus [2], impair the human innate immune system [3] and contribute to the aerosol evaporation [4, 5].

Since the COVID-19 pandemic started, many research groups worldwide have aimed to reveal the relationships between temperature and COVID-19 transmission. Some of these investigated the possibility that the transmissibility is associated with temperature, where the transmissibility is often translated into the number of positive cases [6, 7]; however, these studies did not fully account for the transmission dynamics influenced by PHSMs of various intensities. A few of the earlier studies have explored association between temperature and mortality [6–8] as an indicator of the clinical severity, although they did not address the issue that daily fluctuations in the number of deaths are also vulnerable to the epidemic dynamics. Transmission dynamics of infectious diseases should also be considered when performing the regression models because observation of each case with a contagious disease is not independent, which characteristics is referred to as dependent happening and explicitly distinguishable from other non-communicable diseases; otherwise, such inferences get largely biased [9, 10].

The present study explored the association between temperature and both the transmissibility and the severity of COVID-19 from the early 2020 to the early 2021. We used the effective reproduction number (*R*_*t*_), defined as the mean number of secondary cases generated by a single primary case, to quantify the transmissibility of the ongoing epidemic in Tokyo. To explore the association between temperature and severity, we used case fatality risk (CFR), an epidemiological measurement of severity. Crude CFR calculated from the ratio of the cumulative number of deceased cases to the cumulative number of confirmed cases can underestimate the actual CFR when cases are increasing and overestimate it when they are decreasing due to the time that passes from the onset illness to death. Such issues are also known as right censoring. We therefore estimated the time-delay adjusted CFR for every illness onset date which accounted for the delay.

## Method

### Epidemiological and meteorological data

The data used in the present study were from lab-confirmed, illness onset, and death cases in Tokyo. Meteorological data of temperature, relative humidity, ultraviolet radiation, and wind speed were also analysed.

We used data from 16^th^ February 2020 to analyze the transmissibility with the regression model, as it was the earliest date of the limited publicly available dataset. To analyse severity, data in the regression model were used from 25^th^ May 2020, when the first state of emergency was lifted in Tokyo because CFR may be underestimated given the under-ascertainment rate, and downward ascertained trend early in the epidemic. To avoid the influence of the different infectivity and severity between the previous strain and other evolved strains, e.g., B1.1.7, we cut off the period in both analyses after March 2021.

The daily number of confirmed cases, illness onset cases, and deaths with COVID-19 in Tokyo were collected from 16^th^ January 2020 to 19^th^ March 2021. Confirmed data with age (decades) were also collected from 16^th^ February 2020 to 7^th^ April 2021. To address the measurement of overwhelmed medical situations, we obtained the daily number of cases of emergency transportation whose destination had not been determined within 20 minutes from the start of the Emergency Medical Services team’s request, or who had been refused by at least five medical institutions. To deal with the impact of human mobility, we resorted to Google’s COVID-19 Community Mobility Reports [11], which provides three data-streams on movement in Tokyo: “residual”, “retail and recreation”, and “workplace”. All measures quantify the percentage of deviation from a baseline which indicates the median value for the day of the week during the 5 weeks from 3^rd^ January 2020 to 6^th^ February 2020.

Daily weather data (mean temperature (°C), relative humidity (%), solar radiation as a ultraviolet (MJ/m^2^), and mean wind speed (m/s)) were obtained from the Japan Meteorological Agency including.

### Statistical estimation

The daily *R*_*t*_ estimates were derived from the daily number of confirmed cases and implemented in “EpiNow2” package in R v4.0.2 which method accounted for the week effect and the smoothed renewal process with an appropriate Gaussian process with a squared exponential kernel [12]. The distribution of generation time was adopted from the earlier work [13].

Non-linear and delayed effects of temperature on the transmissibility of COVID-19 were identified by using the generalized additive Gaussian model with the distributed lag non-linear model [14, 15].

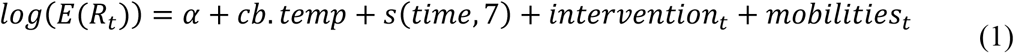

where *cb. temp* represents the nonlinear and delayed exposure-lag-response relationship between the daily *R*_*t*_ and temperature as a form of cross-basis spline function. We used a natural cubic spline with four equally spaced internal knots in the log scale in the cross-basis function [16], accounting for up to 7 days of lag for temperature to examine the lag effect from infection to secondary infection, which is referred to as generation time [13]. Four degrees of freedom (df) of lag were chosen by Akaike Information Criteria (AIC). *s*(.) is a natural cubic spline function. The median value of temperature for calculating relative risk (RR) was 15.3°C. We controlled calendar dates for seasonality (*time*) as a confounder. Seven df per 380 days to *time* were chosen. In addition, *R*_*t*_ would be also influenced by the suppression or mitigation strategies, and other social behavioral changes due to increase in individual awareness of infection [17]. Therefore, we used mobility data, specifically classified into recreation, work, and residual place based on Google mobility data, assuming the three types of places as major possible sites of infection as the variables in the model which involved in some non-pharmaceutical interventions. To compensate above-mentioned issues other than human mobility, we reflected three categorical variables (*intervention*_*t*_) as 0/1/2. *intervention*_*t*_ was imputed as 0 when there were interventions with low intensity on the day *t*, 1 was denoted when the shortened business hours were requested by the Tokyo Metropolitan Government, and 2 was denoted when the state of emergency was declared. Here we did not include a variable for week effect because the framework to estimate *R*_*t*_ has implicitly accounted for the week effect [12]. Distributed lag non-linear model was implemented via “dlnm” package in R v4.0.2.

Subsequently, the association between temperature and the severity of COVID-19 was explored using CFR as a proxy of severity, and the unbiased CFR and daily CFR were estimated [18]. Unbiased CFR was time consistent value while daily CFR was fluctuated on every illness onset date and both accounted for the delay from illness onset to death. We assumed *f*_*s*_ = *F*_*s*_ − *F*_*s*−1_ for *s* > 0 where *F*_*s*_ was cumulative density function of the time-delay. The empirical time-delay distribution was fitted to lognormal, Weibull, gamma, and exponential distributions and best fit gamma distribution with mean 16.6 days and standard deviation 118.4 days by the lowest value of AIC (Supplementary Figure S1). We designated *δ*_*t*_, *d*_*t*_, and *j*_*t*_ as the number of illness onset dates of deaths, deceased dates of deaths, and daily new cases on day *t*, respectively. To adjust for the time-delay, we developed a framework to estimate daily CFR on an illness onset date. Then the time-delay adjusted daily CFR 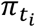 on a time point *t*_*i*_ with observation (*i* = 1,2, …, 299), i.e., from 25^th^ May 2020 to 28^th^ February 2021, was modeled as

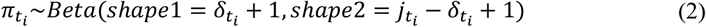

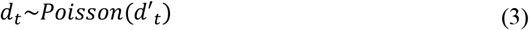

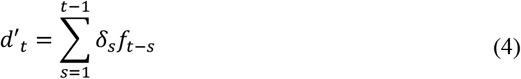

We convoluted *f*_*t*_ with *δ*_*t*_ to obtain the expected number of illness onset dates of deceased cases *d*′_*t*_ and *d*_*t*_ was assumed to follow a Poisson distribution. To deal with the latent variable caused by the convolution, the non-parametric back-projection based on Expectation-Maximization-Smoothing algorithm [19, 20] was conducted by using “surveillance” package in R v4.0.2. The daily CFR was smoothed by beta distribution. In addition, unbiased CFR was estimated as the reference of the daily CFR estimates. *π* denoted the parameter representing the unbiased CFR on the latest day *t*, the likelihood of the estimate *π* was given as

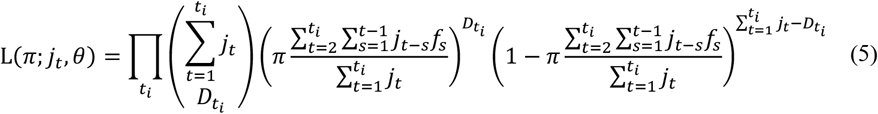

where *t*_*i*_ and 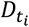 represent and the cumulative number of deaths until reported day *t*_*i*_, respectively [18, 21]. The parameter was estimated using Markov chain Monte Carlo (MCMC) method in a Bayesian framework with the flat prior (*Uniform*(0,1)). We employed Hamiltonian Monte Carlo algorithm with No-U-Turn-Sampler and obtained five chains of 600 thinned samples from 30,000 MCMC iterations where the first 1000 samples of the chains were discarded as burn-in. The MCMC simulations were performed using “rstan” package in R v4.0.2.

We fitted a gamma regression combined with DLNM to estimate the association between temperature and the time-delay adjusted daily CFR 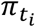 with illness onset dates taking into account the delays in effect of temperature.

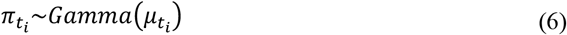

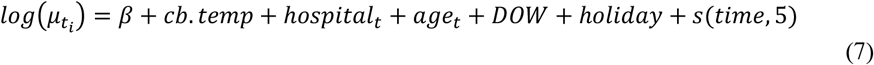

where *cb. temp* represents cross-basis spline function of temperature by a natural cubic spline with four equally spaced internal knots in the log scale in each cross-basis function, accounting for up to 14 days of lag to temperature to examine the period between infection to illness onset, i.e., incubation period which have has previously been explored elsewhere [22]. We considered the 99 % upper bound of incubation period. We also adjusted for the days of the week (*DOW*), holidays (*holiday*), and calendar days for seasonality and long-term trend (*time*) for which five df per 299 days were used by employing a natural cubic spline. *β* is the intercept. The median value of temperature for calculating RR was 18.6°C. Daily age distribution of infected cases with an illness onset day is also critical for CFR as a confounder, i.e., age and age-specific infection fatality risk has an exponential relationship [23]. Because only age distribution with reported dates was publicly available, we back-projected the illness onset date of cases who were over 70 years and in all age groups from the reported dates of cases to calculate the proportion of the daily number of cases over 70 years out of the daily number of cases in all age groups. The time-delay between illness onset to reporting is Weibull distribution and the parameters were adopted from the previous study [24]. In addition, we used the time-series data describing the pressure on medical institutions as *hospital*_*t*_ because whether healthcare system is overloaded or not is a critical factor for CFR.

We conducted sensitivity analysis corresponding to the length of lag and possible meteorological confounders to assess the robustness of the models. As for the lag, the maximum lag day of temperature was set to 5 and 6 to examine the sensitivity of the effect in DLNM for the analysis of transmissibility. For the severity, the maximum lag day of temperature was set to 10 and 12. Regarding meteorological factors as confounders, relative humidity, windspeed, and ultraviolet were included for the analysis of transmissibility, while we considered only relative humidity for the analysis of severity.

## Results

The epidemic curve and estimated median value of *R*_*t*_ with 90% credible intervals (CrI) from 15^th^ February 2020 to 28^th^ February 2021 are shown in Figure 1. Analysing the impact of temperature on *R*_*t*_, the overall cumulative exposure-response relationship of temperature on *R*_*t*_ was non-linear, with lower temperature leading to higher RR (Figure 2. A). The RR corresponding to temperature at the first percentile (3.3°C) was 1.3 (95% confidence interval (CI): 1.1-1.7). Figure 2. B shows the three-dimensional plot of RR with temperature and lags up to 7 days. We found that the greatest risk of cold effects occurs in the day of exposure and risk is increasing in 3-7 days of exposure.

**Figure 1.**
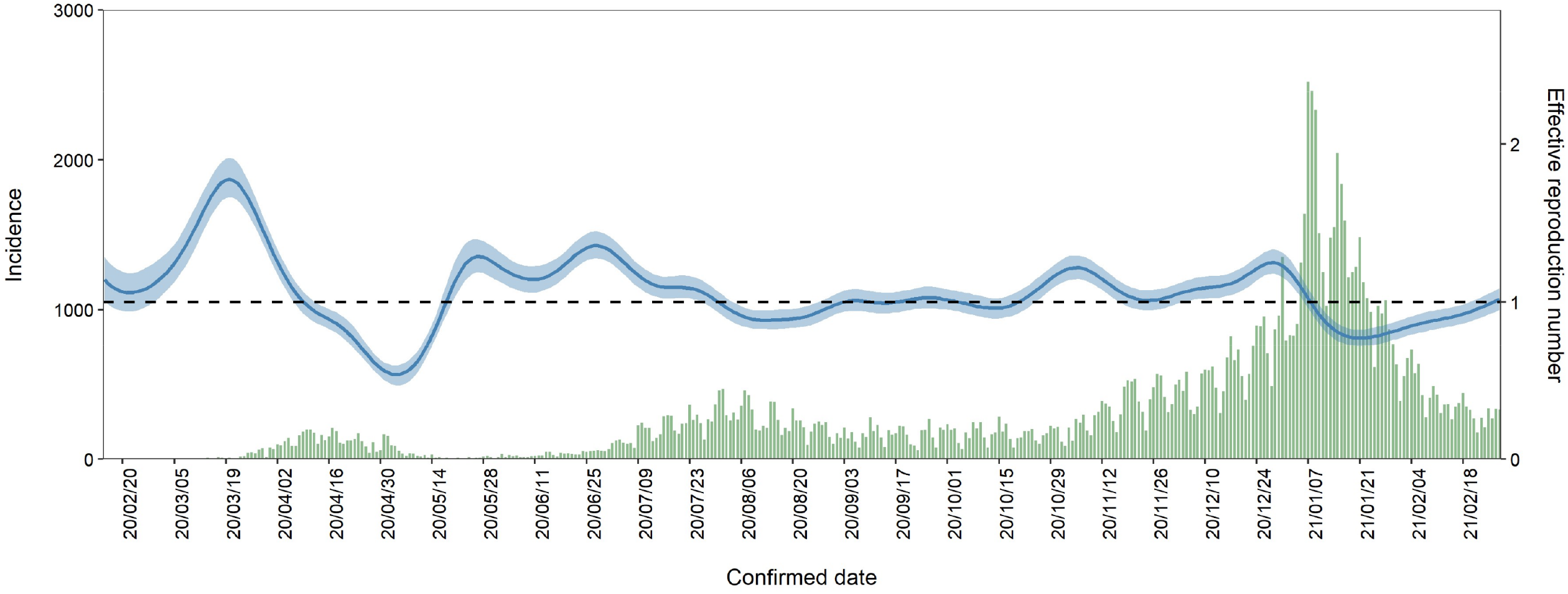
Transmission dynamics from 15^th^ February 2020 to 28^th^ February 2021 in Tokyo, Japan. Blue line represents median, blue shading represents 95% credible intervals of the estimated effective reproduction number from 15^th^ February 2020 to 28^th^ February 2021. Green bars show the observed number of COVID-19 cases with confirmed dates in Tokyo.

**Figure 2.**
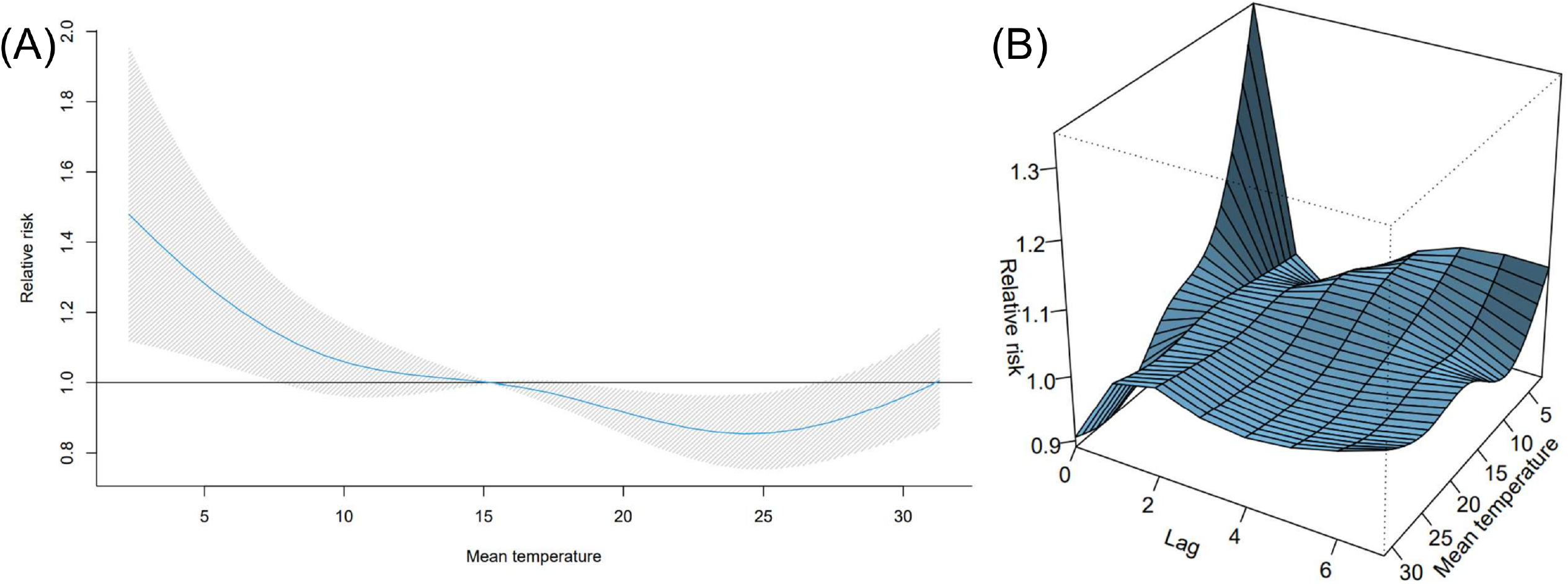
Overall and three-dimensional plots of relative risks with the reference at 15.3°C. (A) The three-dimensional plot of the association between daily mean temperature (°C) and the effective reproduction number over the lags of 7 days. The reference value of temperature was median temperature (15.3°C). (B) The estimated overall effects of mean temperature (°C) over 7 days on *R*_*t*_. Blue line shows the mean relative risks, and 95% confidence intervals are shown in the gray shadings.

Figure 3 shows temporal variation of time-delay adjusted CFR and unbiased CFR from 25^th^ May 2020 to 28^th^ February 2021. As of 28^th^ February 2021, the time-delay adjusted daily CFR and the unbiased CFR were 8.21% (95% CI: 4.50-12.9) and 2.42% (95% CrI: 2.41-2.43), respectively. Figure 3 illustrates the temporal deviations from the baseline value of CFR, i.e., the unbiased CFR. To examine the potential for temperature to contribute to changes in CFR, we estimated overall effect of temperature with the reference of 18.6°C (Figure 4. A). The harmful effect was seen to increase as temperature increased from the reference, and moderate cold temperatures were associated with high RRs of CFR. The three-dimensional plot of RR with temperature and lags for CFR displayed in Figure 4. B, cold temperatures have obvious impact on the day of exposure (lag day 0, the illness onset day) and we found a week delayed effect on both high and cold temperatures.

**Figure 3.**
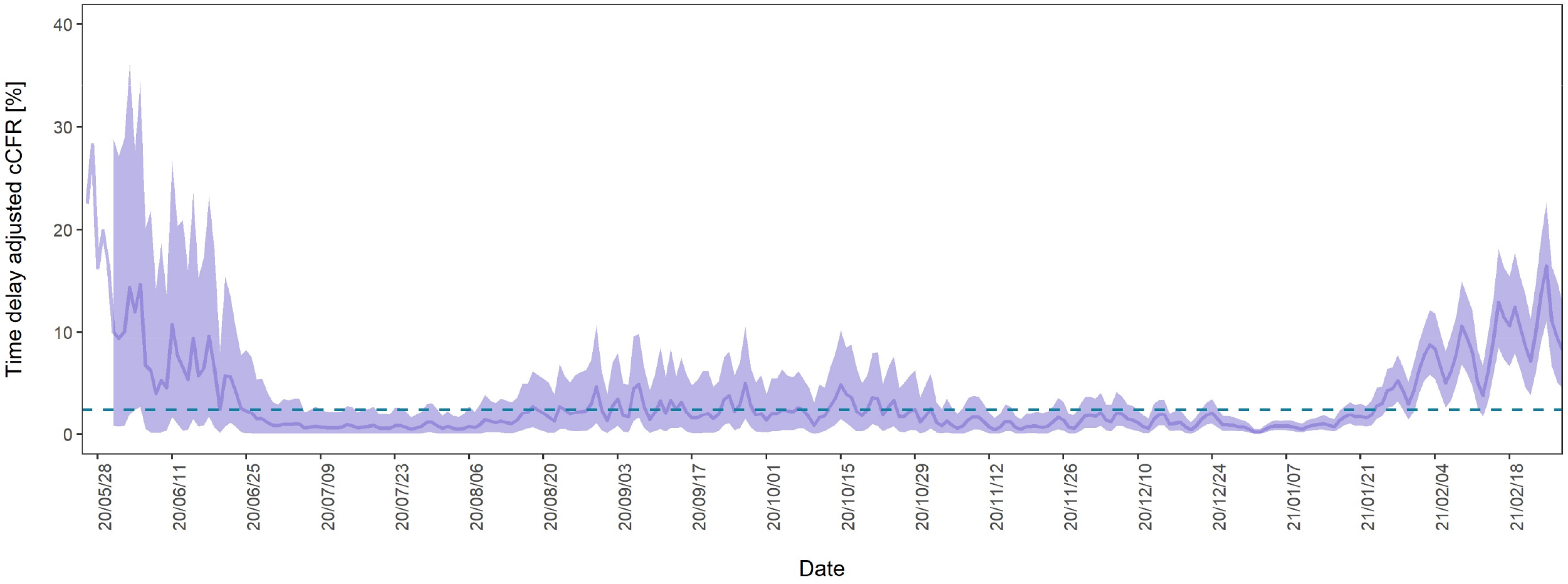
Temporal variation of time-delay adjusted case fatality risks (CFR) with unbiased CFR from 25^th^ May 2020 to 28^th^ February 2021 in Tokyo, Japan. The mean values of time-delay adjusted daily case fatality risks (CFR) from 25^th^ May 2020 to 28^th^ February 2021 are shown with a purple line. The shade region represents the 95% confidence intervals. The blue dot line shows the unbiased case fatality risk as 2.42% (95% credible interval: 2.41-2.43). If the time-delay adjusted daily CFR gets higher or lower, it is caused by random noises or other variables which have causal relationships. The unbiased case fatality risk plays a key role as a reference of the daily CFR.

**Figure 4.**
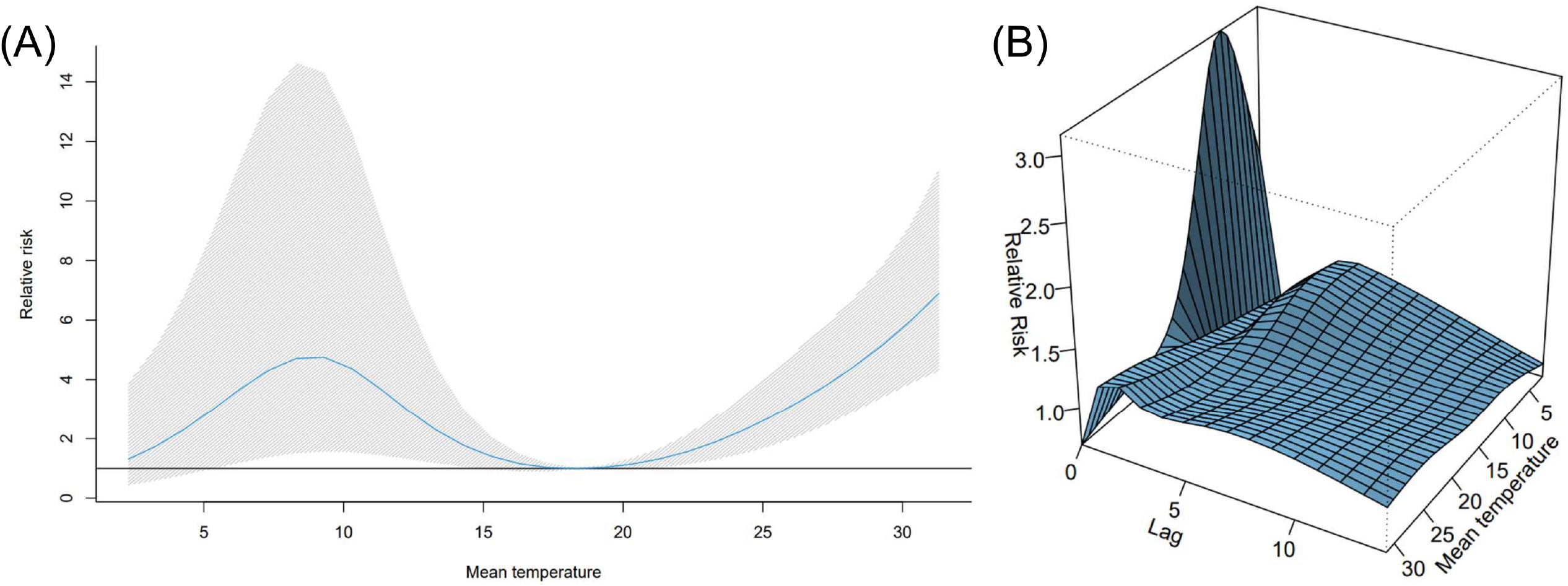
Overall and three-dimensional plots of relative risks with the reference at 18.6°C. (A) The three-dimensional plot of the association between daily mean temperature (°C) and time-delay adjusted case fatality risks (CFR) over the lags of 14 days. The reference value of temperature was median temperature (18.6°C). (B) The estimated overall effects of mean temperature (°C) over 14 days on CFR. Blue line shows the mean relative risks, and 95% confidence intervals are shown in the gray shadings.

Similar results were obtained in sensitivity analysis under different lags and adjustment of several meteorological variables (Supplementary Figure S4-S9) for both of the transmissibility and severity analysis. We assessed the impact on the overall effects and delayed effect and successfully checked the robustness of the primary analysis.

## Discussion

The present study was the first to comprehensively quantify the association between temperature and the epidemiological dynamics of COVID-19 in Tokyo using the effective reproduction number and time-delay adjusted daily CFR. Though the widespread epidemiology of COVID-19 is characterized by the substantial transmissibility and severity which are measured by reproduction number and CFR, there is no study to explore the contribution of temperature using both the rigorous epidemiological measurements appropriately to our best knowledge.

*R*_*t*_ rose explicitly at low temperatures; for example, RR of 3.3°C (1^st^ percentile of temperatures, defined as extreme cold temperature) was estimated as 1.3 (95% CI: 1.1-1.5) in the 0-1 days lag from an infected date (Supplementary Table S1), with median of all the temperatures (15.3°C) as reference temperature. This indicates that the cold effects appear in short lags and overall effect is more plausible in low temperature (Figure 2 A).

The exposure-response relationships of population mobilities and meteorological factors with *R*_*t*_ are consistent with previous works [17, 24, 25]. For example, the residual and workplace mobility change were not slightly related to the fluctuation of *R*_*t*_ while the recreation mobility change was significant. This relationship in Tokyo were reported in the previous study [17]. The exposure-response outcome of solar radiation indicated a significant negative correlation while that of wind speed shows a significant positive correlation (Supplementary Table S1), and similar relationships have been reported elsewhere [24, 25].

As for the severity, we found that low temperatures had a strong association with high CFR in the short lag periods. For example, RR of 2.3°C (1^st^ percentile of temperatures, defined as extreme cold temperature) and 5.8°C (10^th^ percentile of temperatures, defined as extreme cold temperature) were 2.0 (95% CI: 1.2-3.5) and 2.8 (95% CI: 1.7-4.7), respectively, in the 0-2 day lag periods from illness onset dates (Supplementary Table S3), with median of all the temperatures (18.6°C) as reference temperature. While cold effects appear in the short lag period and showed a slight gradual decline, extreme high temperatures were associated with higher CFR from few days after the illness onset, and those effects were stably maintained for two weeks (Supplementary Table S3).

Plausible mechanisms explaining the association between temperature and high CFR of COVID-19 remain undetermined. Even though many studies have postulated seasonal variations and the impact of temperature in transmissibility of infectious diseases, little is known about the association of temperature and severity of contagious disease. Since the common infectious respiratory diseases such as influenza virus, circulate in the cold season, the impact of high temperature on severity is yet to be explored. As a previous study on the impact of heat effect showed, extreme high temperature dampens physiological responses when the body temperature exceeds its normal range [16]. This phenomenon might have contributed to the result observed in the current study. For high CFR of moderate temperature, one of the possible explanations is the difference in human movement which can affect the exposure level with ambient temperature; however, further investigation is needed.

There are several limitations to be noted. First, the assumption for epidemiological time-delays (e.g., generation time and incubation period, etc.) was imposed not to be contracted by meteorological factors or interventions. Second, we did not take into consideration the uncertainty of variables in the regression models, which were derived from the mathematical models. Third, age-specific CFR was not estimated in the present study due to scarce data. However, we attempted to make the best use of the data with back-projected incidence on illness onset dates and controlled the estimated incidence as a confounder in the regression model. Forth, since we used Tokyo data and other geographical locations were not analyzed, the results may vary under different climates. Thus, future studies in multiple locations with our proposed approach are needed. Fifth, the change of ascertainment rate and other confounders (e.g., comorbidity) were not considered in the regression model for CFR. However, we used the empirical data for severity from 25^th^ May 2020 when the state of emergency was lifted, i.e., the end of the first wave of the epidemic in Tokyo in order to avoid higher ascertainment bias and overestimation of CFR.

Despite such limitations, we believe that the present study has provided comprehensive and useful insights on the association between temperature and the characteristics of COVID-19. Higher transmissibility is likely to be seen at low temperatures, while higher severity is likely to present at high and moderately low temperatures. Our findings have important implications to public health responses, as exposure to cold and hot temperatures under a surge of COVID-19 may have the paramount impact on the dynamics and reduction of the burden. We also successfully provided a framework to explore the impact of meteorological factors on the transmission potential and virulence of directly transmitted diseases. Our proposed approach will be applicable for future studies on the relationships between meteorological factors and contagious diseases.

## Data Availability

All data available.

