## Supplementary for "The impact of temperature on the transmission potential and virulence of COVID-19 in Tokyo, Japan"

#### Contents


**1 Daily temperature** **S1**

**2 Time delay distribution from illness onset to death** **S1**

**3 MCMC diagnoses plot** **S2**

**4 Outcomes in the generalized additive models** **S3**
4.1 Analysis of transmissibility . . . S3
4.2 Analysis of severity . . . S3

**5 Sensitivity analysis** **S4**
5.1 Distributed lag nonlinear model for $R_t$ . . . S4
5.2 Distributed lag nonlinear model for CFR . . . S5


### 1 Daily temperature

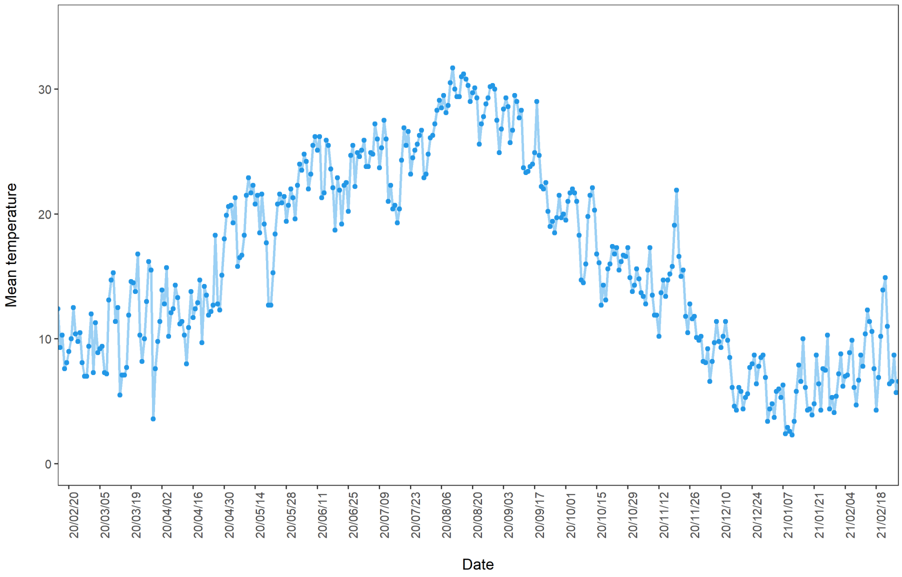


Figure S1: Daily mean temperature from 15th February 2020 to 28th February 2021 in Tokyo, Japan

### 2 Time delay distribution from illness onset to death

We fitted empirical time delay distribution, which was collected from the publicly available data by the Tokyo Metropolitan Government, to Weibull, gamma, lognormal, and exponential distribution. Then gamma distribution was best fit. Figure S1 shows the empirical data and best fitted distribution. The mean and SD are 16.6 days and 118.4 days, respectively.

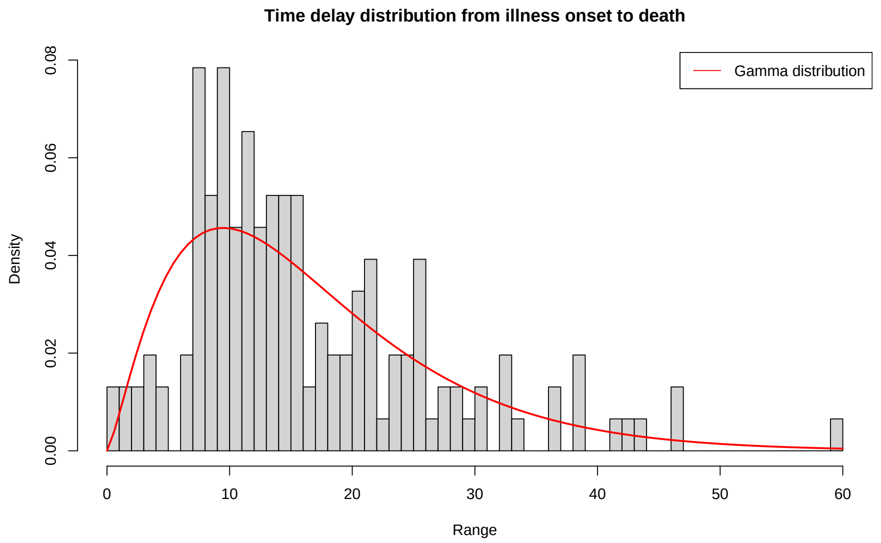


Figure S2: Empirical time delay distribution

### 3 MCMC diagnoses plot

In this section, the convergence of MCMC simulations for Baysian inference are shown in Figure S2. This results indicates that the unbiased CFR is estimated as 2.42% (95%CrI: 2.41-2.43).

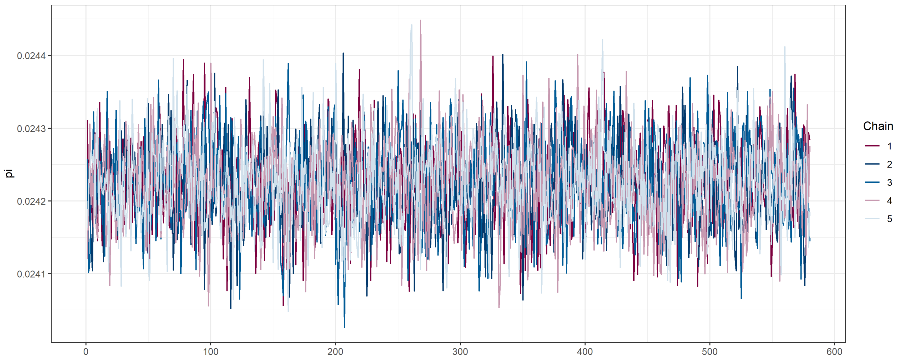


(a) Traceplot

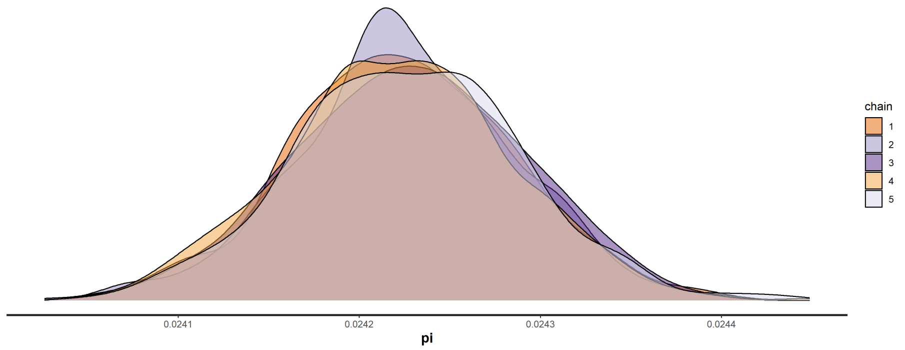


(b) Density plot

Figure S3: Adjustment for maximum lag up to 5 days

### 4 Outcomes in the generalized additive models

#### 4.1 Analysis of transmissibility

Table S1 shows the RR of temperature by 2 days lag which is described in Figure 2. A&B.

Table S1: RR of $R_t$ with stratified lags

| Lag days | Extreme cold | Moderate cold | Moderate hot | Extreme hot |
|---|---|---|---|---|
| 0-1 | 1.3 (1.1-1.5) | 1.1 (1.1-1.2) | 0.89 (0.81-0.98) | 0.9 (0.8-1) |
| 2-3 | 0.93 (0.88-1) | 0.97 (0.93-1) | 1 (0.96-1.1) | 1 (0.96-1.1) |
| 4-5 | 1 (0.98-1.1) | 1 (1-1.1) | 0.98 (0.94-1) | 0.99 (0.94-1.1) |
| 6-7 | 1.1 (1-1.1) | 1 (0.99-1) | 1 (0.97-1) | 1 (0.98-1.1) |

- **Extreme cold**: the first percentile of temperature (3.3 ℃), **Moderate cold**: the 10th percentile of temperature (6.3 ℃), **Moderate hot**: the 90th percentile of temperature (26.9 ℃), **Extreme hot**: the 99th percentile of temperature (30.6 ℃).

Table S3 shows the summary of generalized additive model (GAM).

Table S2: The summary of GAM for $R_t$

| Variable | Estimate | SD | t-value | Pr(>\|t\|) |
|---|---|---|---|---|
| Intercept ($\alpha$) | 0.89 | 0.14 | 6.6 | <0.05 |
| Intervention | -0.15 | 0.018 | -8.3 | <0.05 |
| Residual | 0.0063 | 0.0045 | 1.4 | 0.17 |
| Workplace | 0.003 | 0.0015 | 2 | 0.051 |
| Retail and recreation | 0.012 | 0.0013 | 9 | <0.05 |

#### 4.2 Analysis of severity

Table S3: RR of CFR with stratified lags

| Lag days | Extreme cold | Moderate cold | Moderate hot | Extreme hot |
|---|---|---|---|---|
| 0-2 | 2 (1.2-3.5) | 2.8 (1.7-4.7) | 0.76 (0.53-1.1) | 0.7 (0.44-1.1) |
| 3-5 | 1 (0.85-1.3) | 1.1 (0.9-1.3) | 1.1 (0.97-1.3) | 1.2 (0.98-1.4) |
| 6-8 | 1.2 (1-1.3) | 1.2 (1.1-1.3) | 1.1 (1-1.2) | 1.2 (1.1-1.3) |
| 9-11 | 1 (0.91-1.3) | 1.1 (0.92-1.2) | 1.2 (1.1-1.3) | 1.2 (1.1-1.4) |
| 12-14 | 0.9 (0.8-1) | 0.9 (0.84-1.1) | 1.1 (1-1.2) | 1.1 (1-1.2) |

- **Extreme cold**: the first percentile of temperature (2.8 ℃), **Moderate cold**: the 10th percentile of temperature (5.8 ℃), **Moderate hot**: the 90th percentile of temperature (28.3 ℃), **Extreme hot**: the 99th percentile of temperature (30.8 ℃).

Table S4 & Figure S3 show the summary of GAM.

Table S4: The summary of GAM for CFR

| Variable | Estimate | SD | t-value | Pr(>\|t\|) |
|---|---|---|---|---|
| Intercept ($\beta$) | -1.7 | 0.69 | -2.4 | <0.05 |
| healthcare | 0.0038 | 0.0026 | 1.4 | 0.15 |
| age | -0.34 | 1.1 | -0.32 | 0.75 |
| holiday | -0.18 | 0.14 | -1.3 | 0.21 |
| weekMonday | 0.12 | 0.1 | 1.2 | 0.23 |
| weekSaturday | -0.13 | 0.097 | -1.3 | 0.19 |
| weekSunday | 0.15 | 0.1 | 1.4 | 0.15 |
| weekThursday | 0.18 | 0.098 | 1.8 | 0.07 |
| weekTsuesday | 0.1 | 0.095 | 1.1 | 0.29 |
| weekTuesday | 0.21 | 0.18 | 1.1 | 0.26 |
| weekWednesday | 0.098 | 0.097 | 1 | 0.31 |

### 5 Sensitivity analysis

We conducted sensitivity analysis to transmissibiity and severity by adjusting some possible meteorogical confounders and the length of lags. As to the analysisof transmissibility, Figure S4 shows the results of adjusting meteorogical confounders. Here we considered relative humidity (%), solar radiation as ultraviolet ($MJ/m^2$), and wind speed ($m/s$). Figure S5 and S6 shows the results of changing the length of lags up to 5 and 6 days, respectively.

As for the analysis of severity, we changed the maximum lag days up to 10 and 12 days (Figure S7 and S8). Relative humidity was also controlled in GAM (Figure S8). We chose a linear term for relative humidity because the least value of AIC indicates the 1st df for natural cubic spline was best fit for relative humidity (Figure S9).

#### 5.1 Distributed lag nonlinear model for $R_t$

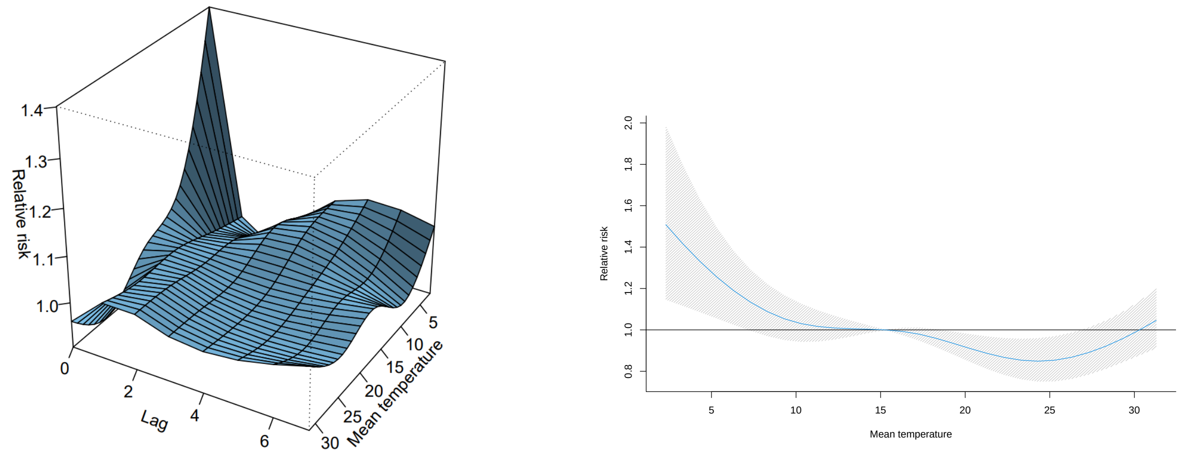


(a) Three-demensional plot

(b) Overall cumulative relationship

Figure S4: Adjustment for maximum lag up to 5 days

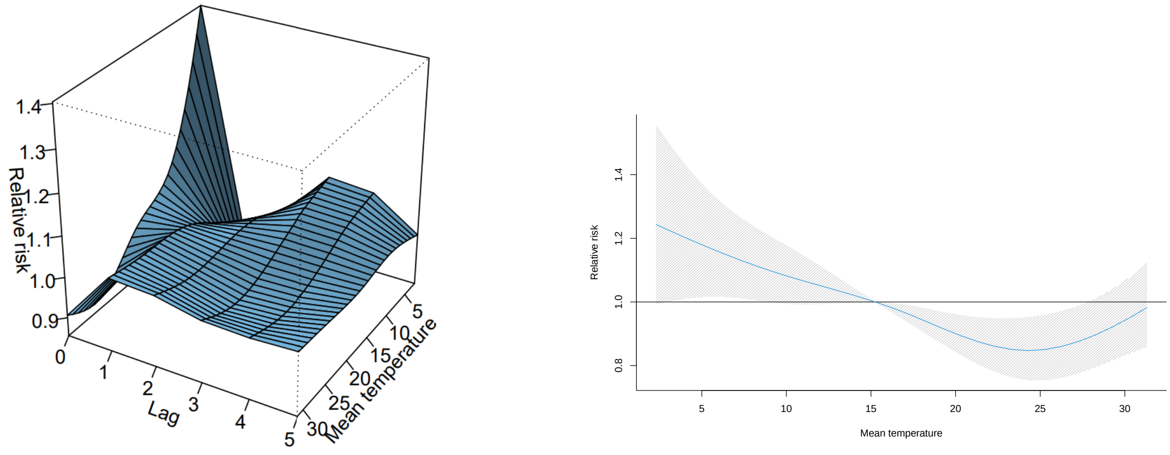


(a) Three-demensional plot (b) Overall cumulative relationship

Figure S5: Adjustment for possible meteorological confounders

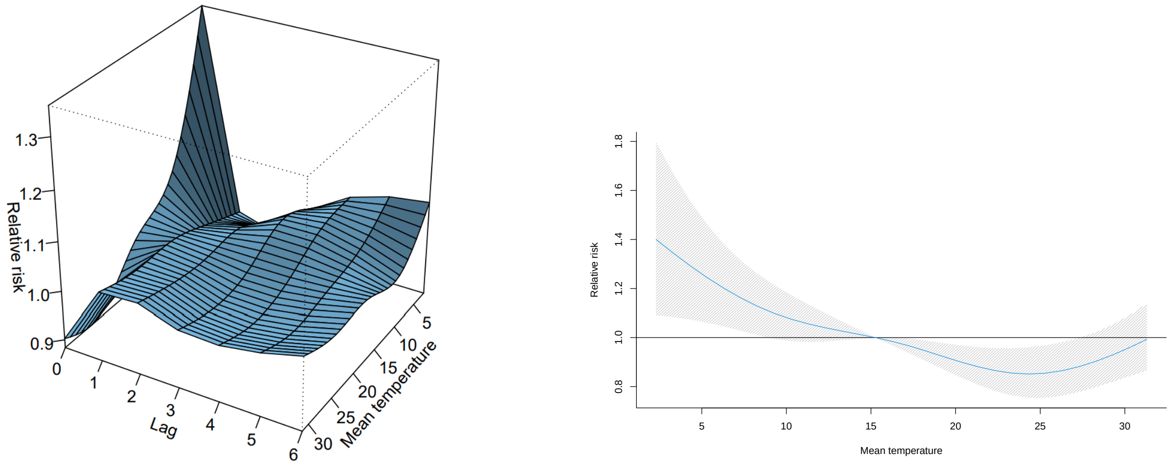


(a) Three-demensional plot (b) Overall cumulative relationship

Figure S6: Adjustment for maximum lags up to 6 days

#### 5.2 Distributed lag nonlinear model for CFR

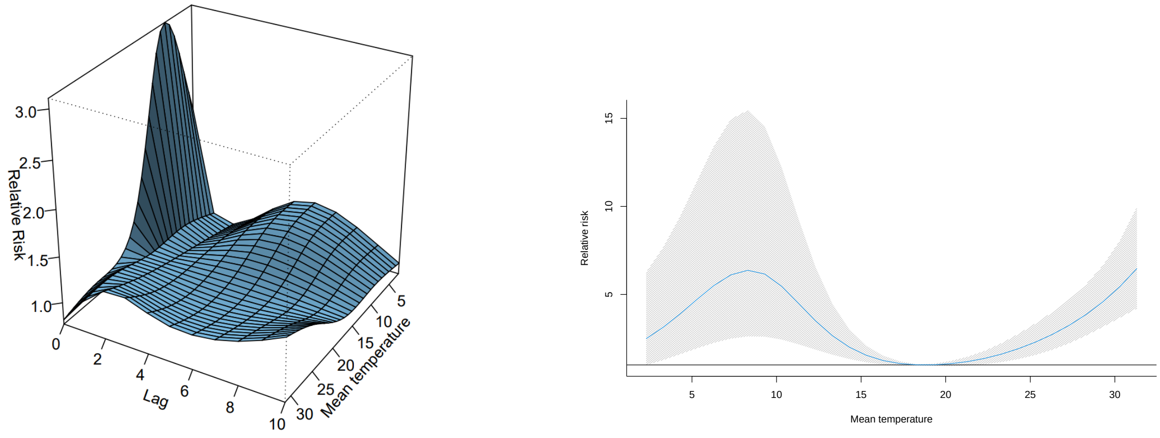


(a) Three-demensional plot (b) Overall cumulative relationship

Figure S7: Adjustment for maximum lag up to 10 days

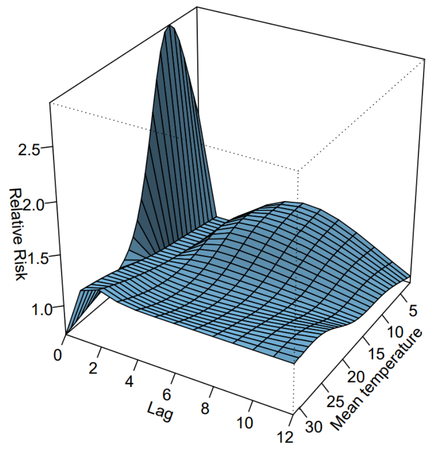


(a) Three-demensional plot

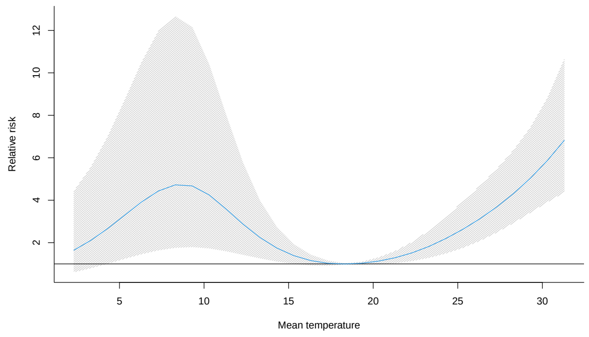


(b) Overall cumulative relationship

Figure S8: Adjustment for maximum lag up to 12 days

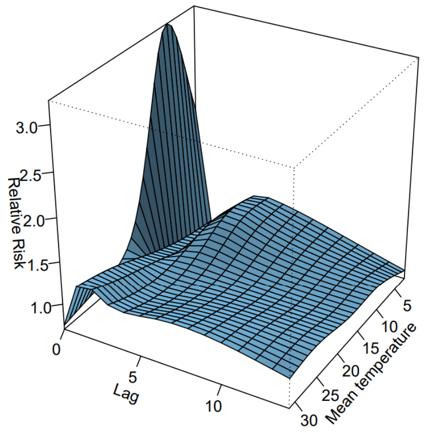


(a) Three-demensional plot

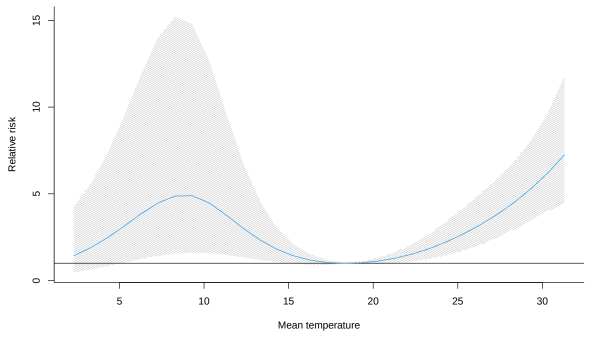


(b) Overall cumulative relationship

Figure S9: Adjustment for relative humidity
